# Antipsychotic Effects on Longitudinal Cognitive Functioning in First-Episode Psychosis: A randomised, triple-blind, placebo-controlled study

**DOI:** 10.1101/2022.02.16.22271103

**Authors:** Kelly Allott, Hok Pan Yuen, Lara Baldwin, Brian O’Donoghue, Alex Fornito, Sidhant Chopra, Barnaby Nelson, Jessica Graham, Melissa J. Kerr, Tina Proffitt, Aswin Ratheesh, Mario Alvarez-Jimenez, Susy Harrigan, Ellie Brown, Andrew D. Thompson, Christos Pantelis, Michael Berk, Patrick D. McGorry, Shona M. Francey, Stephen J. Wood

## Abstract

**Objective:** Cognitive impairment occurs in antipsychotic-naïve first-episode psychosis (FEP), but antipsychotics confound interpretation of the longitudinal course of cognition. The primary aim was to disentangle the effects of illness from antipsychotics on cognition over the first 6-months of FEP treatment.

**Methods:** Randomised, triple-blind placebo-controlled trial (Staged Treatment and Acceptability Guidelines in Early Psychosis; STAGES), where cognition was a secondary outcome. Antipsychotic-naïve FEP patients were allocated to receive risperidone/paliperidone (*N*=38) or placebo (*N*=40) in addition to intensive psychosocial therapy for 6-months. A healthy control group (*N*=42) was also recruited. A cognitive battery assessing attention, working memory, processing speed, verbal fluency, cognitive control and verbal paired-associate learning and memory was administered at baseline and 6-months. Twelve- and 24-month follow-up was also conducted.

**Results:** Over the 6-month trial period, cognitive performance remained stable (working memory, verbal fluency) or improved (attention, processing speed, cognitive control), with no group-by-time interaction evident. The exception was for verbal paired-associate learning and memory, where a significant group-by-time interaction was observed. The placebo and healthy control groups improved, and the medication group deteriorated on immediate paired-associate recall (*p*=0.039) and delayed cued recall (*p*=0.005); effect sizes were medium-to-large. Findings were similar when only trial completers were included in the analysis.

**Conclusions:** Risperidone/paliperidone may cause progression of memory impairment in the early months of FEP. Replication is needed in confirmatory trials. The findings support the need for careful consideration of the risks and benefits of various antipsychotics and the importance of accounting for their cognitive effects in longitudinal research.

**Trial registration:** Australian New Zealand Clinical Trials Registry (http://www.anzctr.org.au/ ACTRN12607000608460).

## Introduction

Widespread cognitive impairments are a core feature of psychotic disorders; being present prior to psychosis onset (1), in medication-naïve patients (2), in first-episode psychosis (FEP) (3), and in unaffected first-degree relatives of probands (4). In both antipsychotic-naïve and antipsychotic-exposed FEP patients, medium-to-large impairments are observed in all assessed cognitive domains. The most severe impairments are in verbal learning and memory, processing speed and working memory (2, 3). Cognitive impairments generally remain quite stable over time (5), although recent longitudinal studies have shown decline in specific cognitive domains many years after FEP (6, 7).

The causes of cognitive change in psychosis remain poorly understood. One contentious factor is the role of antipsychotic medication. It is unclear whether antipsychotics ameliorate, exacerbate, or have negligible effects on cognitive impairments. Clinical trials investigating the effects of atypical antipsychotics on cognitive performance have documented mild improvements in cognition following early acute psychosis (8-10). These reported changes in performance were typically modest (i.e., likely clinically trivial), with further examination suggesting improvements were partly due to cognitive test practice effects (8, 11). Furthermore, small improvements in cognitive functioning following antipsychotic treatment may be due to symptom improvement (9). It is unknown whether cognitive improvements would also be observed with symptom improvement *without* medication. Most trials examining the cognitive effects of antipsychotics have been head-to-head trials without a placebo arm or a healthy control group. Thus, the effect of antipsychotic medication on cognitive functioning cannot be disentangled from the illness itself. As cognitive functioning is one of the most robust predictors of functional recovery following early psychosis (12, 13), it is critical to understand the effects of antipsychotics on cognitive functions.

Most cognitive impairment seems to occur prior to or during the development of full-threshold psychotic illness (14). Because antipsychotics are usually the first line of treatment, it is unclear whether they prevent further deterioration in cognition. One early randomised controlled trial (RCT) showed that, at 2.5-year follow-up, the cognitive functioning of those who experienced an initial 4-week non-medication period (placebo) was no different from those who were immediately prescribed antipsychotics (15). In other words, a 4-week delay in the introduction of antipsychotic medication did not result in long-term harmful effects on cognition. Meta-analyses of the relationship between duration of untreated psychosis (DUP), usually defined as the time between psychotic symptom onset and adequate treatment with antipsychotic medication, and cognitive functioning suggest that the length of DUP is not associated with cognitive performance (16, 17). Moreover, naturalistic studies have shown that higher cumulative use of antipsychotic medications may contribute to poorer cognitive functioning (18, 19), but these findings may be confounded by illness severity and cohort effects. Longitudinal, randomised, placebo-controlled studies of antipsychotic exposure are necessary to determine the effect that initiation and ongoing use of antipsychotic medication has on cognitive function in FEP.

The primary aim of this study was to disentangle the effects of psychotic illness from those of antipsychotic medication on cognitive performance over the first 6-months of treatment for FEP. Therefore, we analysed cognition data collected from a randomised, triple-blind placebo-controlled trial. A healthy control group was also recruited to account for typical cognitive development and test practice effects. A secondary aim was to investigate longer-term changes over 24-month follow-up, which included the period after the 6-month active treatment phase had ended and the prescription of medication became naturalistic. The cognition outcomes reported here are a secondary outcome of the Staged Treatment and Acceptability Guidelines in Early Psychosis (STAGES) trial, where we previously reported that the placebo group had comparable functional outcomes (primary outcome) and clinical outcomes to the group receiving antipsychotic medication (20), and that these treatment groups showed different trajectories of brain volume and function (21, 22).

## Method

### Study Design

STAGES was a randomised, triple-blind, placebo-controlled trial comparing the effects of antipsychotic medication plus intensive psychosocial therapy (medication group) with placebo plus intensive psychosocial therapy (placebo group) in FEP (20, 23). FEP participants were randomised 1:1, stratified by sex and DUP, within six permuted blocks. DUP was included as a three-level factor (0–30, 31–90, >90 days). Clinicians, patients, and research assistants conducting the assessments remained blind to treatment allocation throughout the trial. The active treatment phase was 6-months; between the 6- and 24-month follow-up patients in either group received antipsychotic medication and psychosocial treatments as determined by their practitioner. To account for practice effects, a healthy control group who received no treatment was also recruited. Assessments for the FEP group occurred at baseline (prior to treatment), 6-, 12- and 24-months, whereas healthy controls were only assessed at baseline and 6-months. The Melbourne Health Human Research Ethics Committee approved the study (MH-HREC: #2007:616). Trial registration was with the Australian New Zealand Clinical Trials Registry (ACTRN12607000608460).

### Participants

FEP patients were aged 15-25 years and presented for treatment with the Early Psychosis Prevention and Intervention Centre, a specialist public early psychosis program, which treats young people with FEP in a catchment area covering Melbourne’s northern and western suburbs, Australia. Participants met criteria for a DSM-IV psychotic disorder, including schizophrenia, schizophreniform disorder, schizoaffective disorder, delusional disorder, depression with psychosis, substance-induced psychotic disorder, or psychosis not otherwise specified, confirmed using the Structured Clinical Interview for DSM-IV for Axis I disorders. Inclusion criteria were: ability to provide informed consent; English language comprehension; DUP <6 months; low past exposure to antipsychotic medication (<7 days or lifetime maximum 1750mg chlorpromazine equivalent); no previous treatment with lithium or anticonvulsant medication; and to minimise risk: living in stable accommodation; low risk to self/others (20, 23).

Healthy controls that matched FEP participants on age, sex, and socioeconomic status were recruited via flyers, snowballing, and social media advertisements. Inclusion criteria for healthy controls were: aged 15-25 years and English language comprehension. Exclusion criteria included: history of psychotic disorder or any current mental disorder (screened using the Structured Clinical Interview for DSM-IV Axis I Disorders, Research Version, Non-Patient Edition); head injury or neurological disorder; and studied psychology at university (to mitigate exposure to cognitive tests). All participants gave written informed consent, including parent/guardian consent for participants <18 years. All participants were reimbursed $50AUD per assessment.

### Treatment

FEP patients received weekly cognitive-behavioural case management, a comprehensive manualised formulation-based psychological and psychoeducation treatment for early psychosis (20). Patients also saw a psychiatrist weekly and received close monitoring, family therapy, vocational support, and 24-hour crisis response and outreach as required. Patients randomised to the medication group received either 1mg risperidone (*n*=34) or 3mg paliperidone (*n*=4), depending on the availability of medication and matched placebo (20). The starting dose was gradually increased according to clinical response by the blinded study doctor. The same procedure was followed for patients randomised to the placebo group, who received a placebo pill that was identical to the active medication in taste, appearance, and packaging. To ensure participant safety, strict study medication discontinuation criteria were applied in situations of increased risk to self/others or worsening mental state/ functioning and an alternative open-label antipsychotic medication was offered (23).

### Cognitive Battery

The Information and Picture Completion subtests of the Wechsler Adult Intelligence Scale–3^rd^ Edition (WAIS-III) were administered at baseline to estimate current IQ. The repeated cognitive battery included measures of attention (WAIS-III Digit Span: forward), working memory (WAIS-III Digit Span: backward), processing speed (WAIS-III Digit Symbol-Coding), verbal fluency (Controlled Oral Word Association Test and animal fluency), processing speed and inhibition (Golden Stroop: words, colours and colour-word subtests), and verbal learning and memory (immediate paired-associate recall, total paired-associate learning: trials 1-3, and delayed cued recall from the Melbourne Paired-Associate Learning task (24); A/B forms were alternated at each time-point). Raw scores were used in the analysis.

### Statistical Analysis

Comparison of the three groups on baseline measurements was conducted using analysis of variance (ANOVA), followed by pairwise comparison using Fisher’s least significant difference (LSD) test. To assess rate of change in cognitive performance, group comparisons on each cognitive outcome were conducted using linear mixed-effects (LME) modelling analysis with random effects for intercept and slope and group-by-time interaction. LME analysis includes all cases with data on at least one time-point. The actual times of assessment (i.e., total weeks from baseline to 6-month cognitive assessment) were used in the analysis. For significant group-by-time interactions, pairwise comparisons were conducted with effect size indexed using partial eta squared (*η*_*p*_^2^), which indicates the proportion of variance explained by a given term in a model after accounting for variance explained by the other terms in the model, whereby *η*_*p*_^2^=0.01 is considered small, 0.06 medium and 0.14 a large effect (25). For the primary aim and analysis, we compared the three groups over the 6-month trial period. For the secondary aim and analyses, only the two FEP groups (placebo/medication) were compared over 24-months. In accordance with CONSORT recommendations (26), a per protocol LME analysis was also performed, i.e., only including participants who remained on their allocated trial medication (placebo/medication) throughout the 6-month trial period. A significance level of *p*<0.05 was used. Adjustment for multiple testing was not applied as this study was considered exploratory (not confirmatory) and was not specifically powered for the cognition outcomes. In this scenario, priority is given to minimising type II error and generating evidence to inform future hypothesis-driven studies (27, 28). The analysis was conducted using the *nlme* and *effectsize R* packages (25, 29).

## Results

Figure 1 shows the participant flow into the study. Demographic and clinical characteristics of each group are presented in Table 1. The three groups did not significantly differ in mean age or sex distribution. The placebo and medication groups did not significantly differ on any baseline variable, including cognitive functioning. As expected, healthy controls had a significantly higher mean education, IQ, social and occupational functioning and lower symptomatology than the FEP groups. Healthy controls also had higher mean performance than the FEP groups on all cognitive measures at baseline, which was significant apart from Digit Span: Forward, and Stroop: Words (Supplementary Table 1).

**Figure 1.**
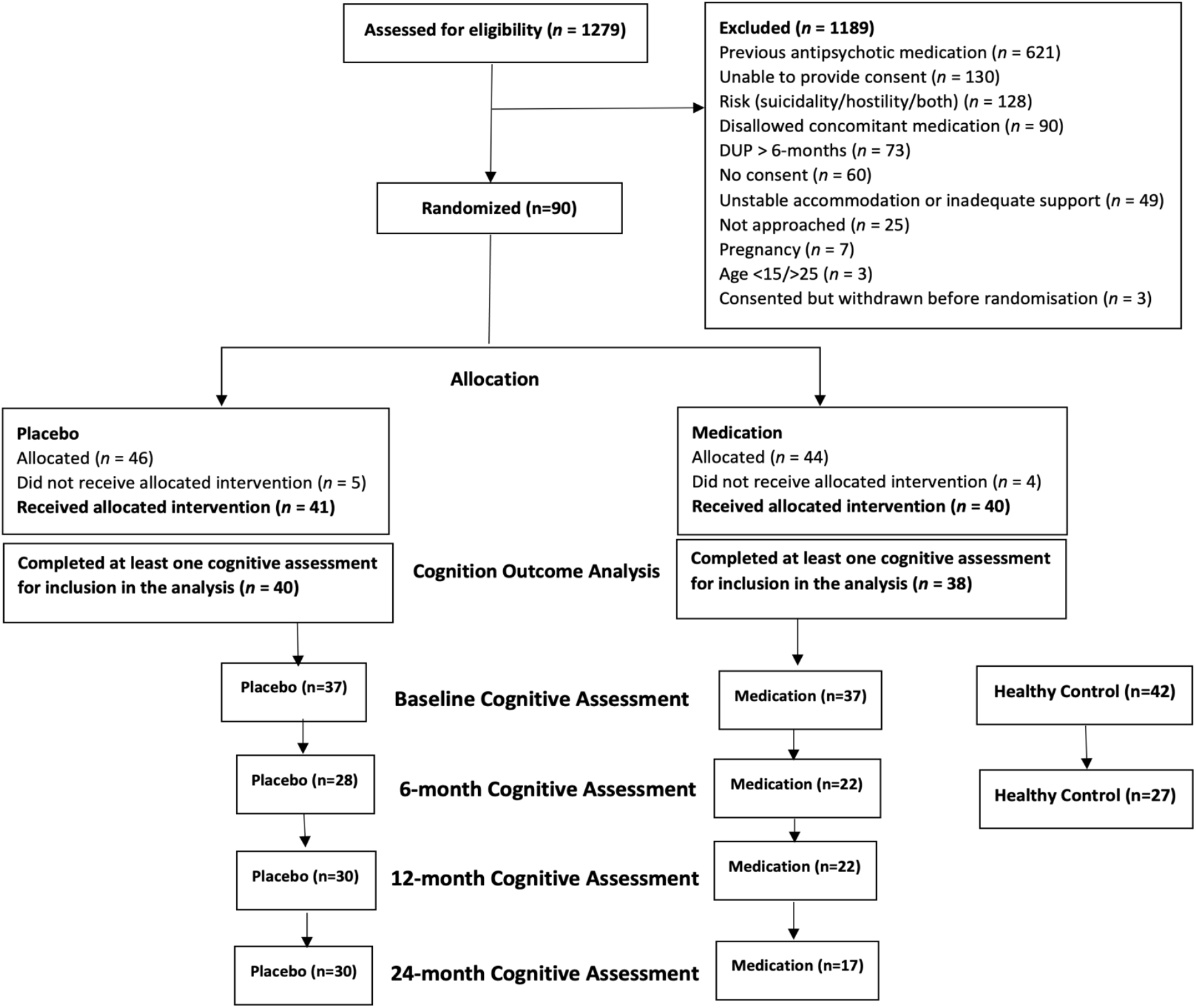
Participant flow

**Table 1.** Participant characteristics at baseline

|  | First-episode psychosis |  | Healthy Control<br>(N=42) |
| --- | --- | --- | --- |
|  | Placebo<br>(N=40) | Medication<br>(N=38) |  |
| Age, years (SD) | 18.2 (2.7) | 18.7 (2.8) | 19.2 (3.0) |
| Females, N (%) | 23 (57.5%) | 21 (55.3%) | 28 (66.7%) |
| Education, years (SD) | 11.8 (1.8) | 12.0 (3.0) | 13.4 (2.2) |
| Estimated IQ | 90.5 (13.4) | 91.8 (14.3) | 107.6 (10.0) |
| BPRS Total, mean (SD) | 58.6 (9.0) | 57.5 (9.9) | 31.6 (3.6) |
| BPRS Psychotic, mean (SD) | 15.0 (3.1) | 13.9 (3.7) | 4.1 (0.3) |
| SOFAS, mean (SD) | 54.1 (13.1) | 53.7 (10.9) | 80.5 (8.9) |
| SANS Total, mean (SD) | 27.0 (15.0) | 25.9 (14.8) | - |
| DUP, N (%) |  |  |  |
| 0-30 days | 6 (15.0%) | 6 (15.8%) | - |
| 31-90 days | 14 (35.0%) | 12 (31.6%) | - |
| >90 days | 20 (50.0%) | 20 (52.6%) | - |
| Psychotic Diagnosis, N (%) |  |  |  |
| Major depression with psychosis | 8 (20.0%) | 8 (21.1%) | - |
| Schizophreniform disorder | 6 (15.0%) | 9 (23.7%) | - |
| Psychotic disorder NOS | 12 (30.0%) | 8 (21.1%) | - |
| Schizophrenia | 7 (17.5%) | 5 (13.2%) | - |
| Substance-induced psychotic disorder | 6 (15.0%) | 4 (10.5%) | - |
| Delusional disorder | 1 (2.5%) | 4 (10.5%) | - |

BPRS=Brief Psychiatric Rating Scale; SOFAS=Social and Occupational Functioning Assessment Scale; SANS=Scale for the Assessment of Negative Symptoms; DUP=duration of untreated psychosis; NOS=not otherwise specified.

### Primary analysis: 6-month RCT phase

The three groups were compared in terms of rate of change in cognitive functioning from baseline to 6-months using LME analyses (Table 2). A significant effect of time was observed on tasks of attention, processing speed and cognitive control (i.e., Digit Span: forward, Stroop: colour, word and colour-word, and Digit Symbol-Coding); the estimated rates of change were all positive, indicating a significant overall increase from baseline to 6-months. Stable cognitive performance was observed in working memory (i.e., Digit Span: backward) and verbal fluency (letter/animal). There were no group-by-time interactions on any of these measures.

**Table 2.** Results of LME analysis comparing the three groups on the rate of cognitive change from baseline to 6-months

|  | P-values |  |  | Estimated rate of change |  |  |
| --- | --- | --- | --- | --- | --- | --- |
|  | Group x time interaction | Group | Time | Coefficient | Standard Error | n |
| Digit Span: forward | 0.152 | 0.054 | **0.001** | 0.022 | 0.006 | 118 |
| Digit Span: backward | 0.354 | **<0.001** | 0.683 | 0.002 | 0.007 | 118 |
| Digit Symbol-Coding | 0.114 | **<0.001** | **<0.001** | 0.185 | 0.043 | 107 |
| Stroop: words | 0.336 | **0.042** | **0.005** | 0.115 | 0.039 | 116 |
| Stroop: colours | 0.237 | **0.001** | **0.007** | 0.096 | 0.035 | 116 |
| Stroop: colour-word | 0.968 | **<0.001** | **0.006** | 0.082 | 0.029 | 116 |
| Letter fluency | 0.187 | **0.001** | 0.204 | 0.040 | 0.025 | 117 |
| Animal fluency | 0.978 | **0.010** | 0.978 | 0.0001 | 0.019 | 116 |
| Immediate paired-associate recall (Trial 1) | **0.039** | **<0.001** | 0.090 | 0.015 | 0.009 | 118 |
| Total paired-associate learning (Trials 1-3) | 0.068 | **<0.001** | 0.277 | 0.021 | 0.019 | 118 |
| Delayed cued recall | **0.005** | **<0.001** | >0.999 | -0.0001 | 0.007 | 117 |

Significant group-by-time interactions were found on the verbal paired-associate learning and memory task, specifically immediate paired-associate recall (Trial 1) (*p*=0.039) and delayed cued recall (*p*=0.005). Total paired-associate learning (Trials 1-3) showed a similar, though non-significant (*p*=0.068) interaction effect (Table 2). Figure 2 plots the estimated trends from baseline to 6-months for the three groups on these three measures (Supplementary Figure 1 plots remaining measures). Figure 2a illustrates the difference in rate of change in immediate paired-associate recall (Trial 1), where the placebo and healthy control groups show an improvement over time and the medication group shows a deterioration. Similar patterns can be observed in Figures 2b and 2c. Pairwise comparisons showed that there was a significant decline in the medication group compared to the healthy control (*p*=0.023; *η*_*p*_^2^=0.071) and placebo (*p*=0.037; *η*_*p*_^2^=0.060) groups for immediate paired-associate recall, as well as total paired-associate learning (healthy control vs. medication *p*=0.039; *η*_*p*_^2^=0.059; placebo vs. medication *p*=0.020; *η*_*p*_^2^=0.074). For delayed cued recall, the medication group again showed significant decline when compared to the placebo group (*p*=0.001; *η*_*p*_^2^=0.137); however, neither group showed a significant difference in rate of change when compared to the healthy group (healthy control vs. placebo *p*=0.060; *η*_*p*_^2^=0.049; healthy control vs. medication *p*=0.098; *η*_*p*_^2^=0.038).

**Figure 2.**
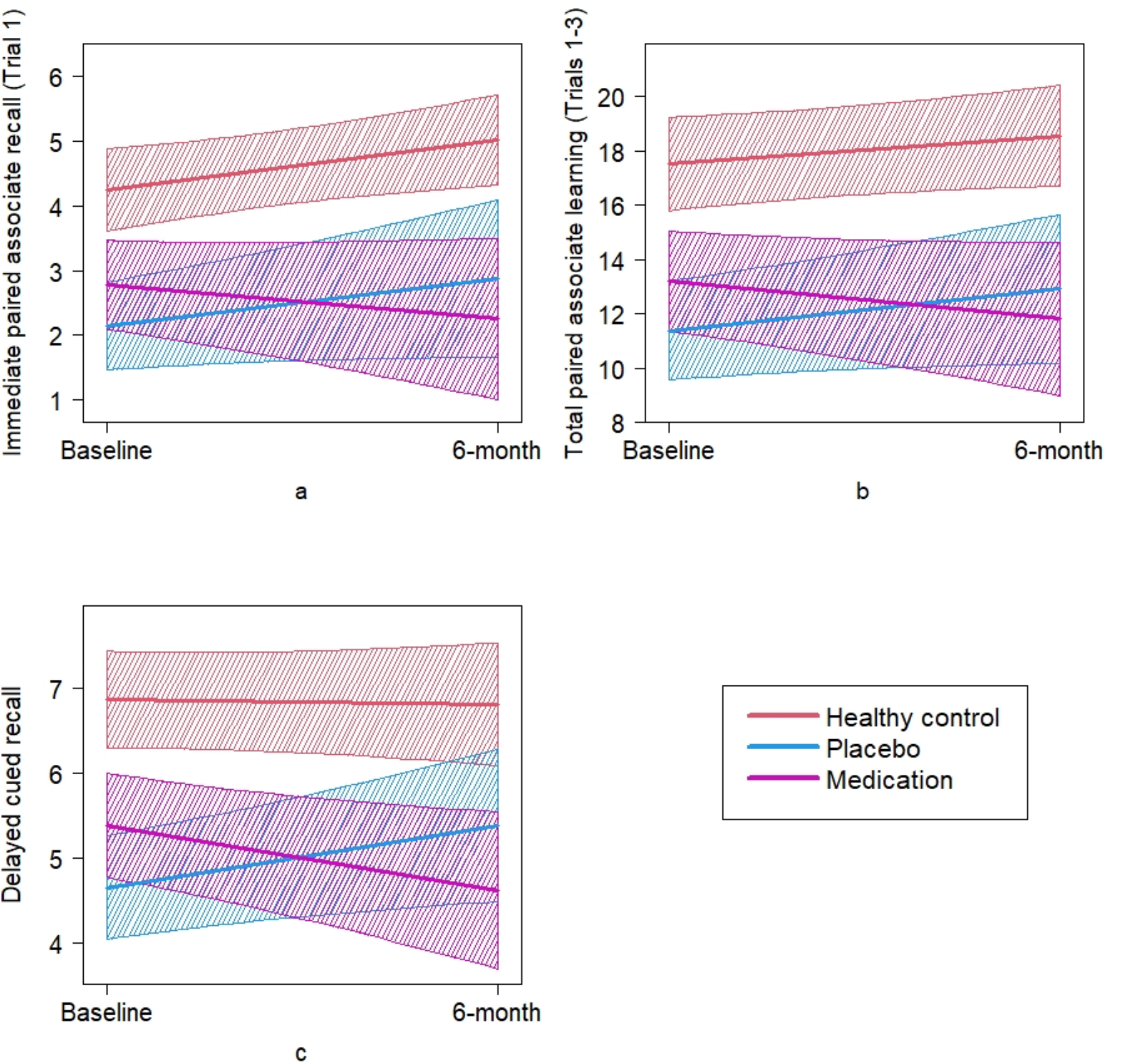
Plots of estimated trends from baseline to 6-months and associated 95% confidence bands for verbal paired associate learning and memory.

During the 6-month trial period, the mean cumulative dose of antipsychotics (olanzapine equivalent mg) was 897.6 (SD=484.9; range 330-2105) in the medication group and 351.7 (SD=507.9; range 0-1803) in the placebo group, which was significantly different (*p*<0.001). Twelve patients from the placebo and 11 from the medication group discontinued trial medication and were prescribed an alternative medication (e.g., antipsychotic); thus, the placebo group was not a pure placebo group and medication group was not a pure risperidone/paliperidone group. To test the robustness of the intention-to-treat findings, the analysis was repeated including only per protocol trial completers (placebo n=16, medication n=11, healthy control n=42; Supplementary Table 2; Supplementary Figure 2). The results remained similar, with significant group-by-time interactions on the verbal paired-associate learning task, specifically total paired-associate learning (Trials 1-3) (*p*=0.035) and delayed cued recall (*p*<0.001). Immediate paired-associate recall (Trial 1) was non-significant (*p*=0.059), but the interaction effect seen was similar to that in the full sample. Pairwise comparisons showed that all three groups differed from each other in the rate of change for delayed cued recall (healthy control vs. placebo *p*=0.009; *η*_*p*_^2^=0.137; healthy control vs. medication *p*=0.011; *η*_*p*_^2^=0.130; placebo vs. medication *p*<0.001; *η*_*p*_^2^=0.290). There was a significant difference in the rate of change between the medication group and the other two groups for immediate paired-associate recall (healthy control vs. medication *p*=0.034; *η*_*p*_^2^=0.092; placebo vs. medication *p*=0.036; *η*_*p*_^2^=0.090) and total paired-associate learning (healthy control vs. medication *p*=0.020; *η*_*p*_^2^=0.110; placebo vs. medication *p*=0.015; *η*_*p*_^2^=0.120). Additionally, a group-by-time interaction was found for digit symbol-coding (*p*=0.016), where the healthy control group improved, but the medication and placebo groups remained stable (healthy control vs. placebo *p*=0.028; *η*_*p*_^2^=0.099; healthy control vs. medication *p*=0.019; *η*_*p*_^2^=0.111).

### Secondary analysis: 24-month uncontrolled follow-up phase

The placebo and medication groups were compared on the rate of change in cognitive functioning from baseline to 24-months using LME modelling (Supplementary Table 3; Supplementary Figure 3). Testing for quadratic trend produced non-significant results and therefore linear trend was used. For almost all cognitive measures (except animal fluency) there was a significant effect of time, where the estimated rates of change were all positive, indicating a significant improvement in cognitive functioning overall from baseline to 24-months. One significant group-by-time interaction was observed for the Stroop: colours task (Figure 3), indicating that the placebo group showed a smaller rate of improvement than the medication group (*p*=0.032; *η*_*p*_^2^=0.031).

**Figure 3.**
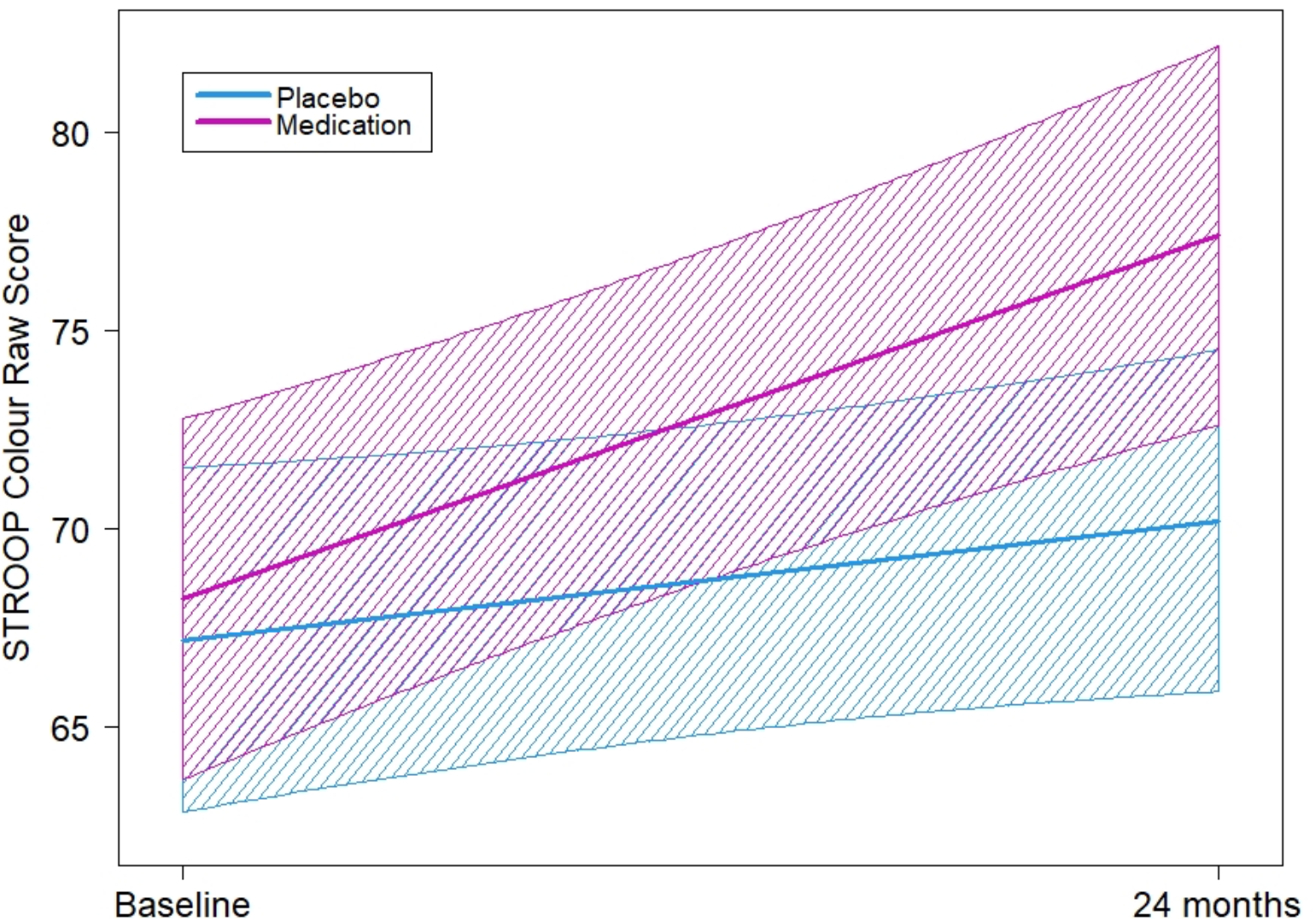
Plot of estimated trends from baseline to 24-months and associated 95% confidence bands for the Stroop: Colour task.

We repeated the analysis by only including trial completers (Supplementary Table 4). Again, there was a significant effect of time for several measures (Digit Span: forward and backward, Digit Symbol-Coding, Stroop: colour and colour-word), where the estimated rates of change were positive, indicating a significant improvement in cognitive functioning overall from baseline to 24-months. There were no significant group-by-time interactions.

## Discussion

This randomised, triple-blind, placebo-controlled study enabled the longitudinal (6-month) cognitive effects of psychotic illness to be disentangled from those of antipsychotic medication. The primary finding was that exposure to risperidone/paliperidone over the 6-month controlled trial phase was associated with a decline in verbal paired-associate learning and memory. In contrast, patients who received placebo improved in verbal paired-associate learning and memory at a similar rate to healthy controls. The effects were medium-to-large. This finding remained when only trial completers were included in the analysis. A clear strength of this challenging study was the placebo-controlled randomised design, which mitigates bias, ensures participant comparability from the outset, and allows determination of causality. Importantly, the medication and placebo groups did not significantly differ at baseline on any cognitive measure, length of DUP, or other illness variables. Over the 24-month period, the findings regarding verbal paired-associate learning and memory were no longer evident and the risperidone/paliperidone group showed a greater rate of improvement on a visual processing speed task than the placebo group; this finding was not upheld when only trial completers were included in the analysis. The 24-month follow-up findings should be interpreted cautiously because this included an 18-month uncontrolled period where the prescription of medication became naturalistic.

A significant implication of the findings is that, at least for some patients, partially or completely withholding antipsychotics (when clinically safe to do so) is not harmful to cognitive functioning and may be beneficial for verbal paired-associate learning and memory in the early course of FEP. Randomly lengthening the DUP (in the placebo group), customarily defined as the onset of psychotic symptoms to first antipsychotic treatment, was not associated with a worsening of cognition in any domain measured. This finding is consistent with previous meta-analyses showing a negligible relationship between DUP and cognitive function (16, 17). On most cognitive measures, the change in cognition was similar across the placebo, medication and healthy control groups, suggesting improvements over 6-months are likely explained by practice effects (11).

A second key implication of the findings is that risperidone/paliperidone may cause progression of memory impairment, at least for some patients with FEP. Over 6-months of treatment, the performance of the medication group declined below baseline (pre-treatment) performance. These effects of risperidone/paliperidone may arise because of their antagonism at dopamine (D_2_) receptors, with previous research showing a significant correlation between risperidone dose and extrastriatal D_2/3_ occupancy (30). Uncontrolled studies involving antipsychotic-naïve first-episode patients have shown risperidone was associated with decline in spatial working memory over 6-weeks (31), and attention and planning time over 12-weeks (30), with a significant negative correlation observed between extrastriatal D_2/3_ occupancy and cognitive performance in the latter study. While these specific cognitive functions were not assessed in the current study, D_2_ receptors are prominently expressed in brain regions involved in paired-associate learning and memory, particularly the striatum, prefrontal cortex and hippocampus (32, 33). Acute administration of risperidone into the hippocampus of rats decreases local cerebral glucose utilisation (34), suggesting that D_2_ antagonists, such as risperidone, may impair the ability to encode and consolidate new information. Furthermore, the paired-associate learning task was the most effortful and lengthy of all tasks in the cognitive battery, and it is possible that D_2_ antagonism impaired the sustained cognitive effort required for this task more than others. Importantly, given antipsychotics as a group differ greatly in their off-target effects outside of dopamine (D_2_) antagonism, and may differ in effects on cognition as well (35), this study cannot be generalised to all antipsychotics.

While an anticholinergic mechanism could plausibly explain the observed decline in verbal learning and memory (36-38), risperidone/paliperidone have a relatively low risk for anticholinergic adverse effects (39). Furthermore, the doses prescribed here, while still effective (20), were lower than these previous studies. Use of other medications, such as antidepressants and benzodiazepines also have low anticholinergic activity (39) and did not differ between the treatment groups (20). The anticholinergic benztropine was prescribed PRN to six patients (n=1 placebo, n=5 medication group); only three (8% of medication group) received benztropine during the 6-month trial period; all six were withdrawn cases and prescribed benztropine after withdrawal and were excluded from the completer analysis. Thus, anticholinergic effects are an unlikely explanation for the findings.

The specificity of the negative effect of risperidone/paliperidone to learning and memory is notable given that impairment in this cognitive domain is particularly severe at the first-episode (3), is associated with poor community/vocational and social functioning (12), and shows decline in longitudinal studies (although not always in the verbal domain) (6, 7). While the cognitive effects of medication cannot be accurately determined in naturalistic longitudinal cohort studies since cumulative antipsychotic exposure is confounded with the effect of severity of illness, the current findings suggest antipsychotic medication (in this case, risperidone/paliperidone) may contribute to the decline in learning/memory observed in these studies. In support of this, antipsychotic dose reduction has been associated with improvements in cognitive functioning (including learning and memory) in both naturalistic longitudinal studies (18) and RCTs (40) involving FEP patients. In the current study, a relatively small sample size and the naturalistic nature of treatment post-6-months may explain the wash-out in group differences over 24-months.

It is important to acknowledge that this was a highly selected FEP sample due to the specific inclusion/exclusion criteria of the trial, so findings may not generalise to all FEP individuals and certainly require replication in confirmatory trials. Nevertheless, the degree of cognitive impairment in the sample at baseline was medium-to-large, which is comparable to other FEP samples (3). This confirms that cognitive impairment is a central feature of psychosis that is independent of the effects of treatment, including antipsychotics. Another limitation is that a high percentage of participants did not complete the trial as planned. However, the completer analysis was consistent with the primary analysis, suggesting that the results are robust. Finally, the lack of healthy control cognitive data beyond 6-months limits the interpretation of the cognitive trajectories of the FEP groups over the longer-term follow-up.

In conclusion, the findings highlight the importance of accounting for the cognitive effects of antipsychotic medication in FEP. Careful consideration of the risks and benefits of antipsychotic initiation and maintenance is critical and must occur within a shared decision-making framework. Indeed, a recent study showed that of various antipsychotic side-effects, memory impairment had the strongest influence on the medication preferences of people with schizophrenia (41). Future research must investigate within-class differences in the effects of antipsychotics on cognition. If this confirms what is reported here, then efforts to address cognitive impairment in FEP should consider medication effects, including differential cognitive profiles (35), alongside behavioural treatments, such as cognitive remediation or compensation.

## Data Availability

All data produced in the present study are available upon reasonable request to the authors.

## Acknowledgements

The study was supported by Jannsen-Cilag, Australia and the National Health and Medical Research Council, Australia (#1064704).

## Authorship

Dr Allott affirms that she has access to all data from the study, both what is reported and what is unreported, and also that she had complete freedom to direct its analysis and its reporting, without influence from the sponsors. The corresponding author also affirms that there was no editorial direction or censorship from the trial sponsors.

## Disclosures/Conflict of Interest

KA is supported by a NHMRC Career Development Fellowship (1141207) and a University of Melbourne Dame Kate Campbell Fellowship. MB is supported by a NHMRC Senior Principal Research Fellowship (1156072). MB has received Grant/Research Support from the NIH, Cooperative Research Centre, Simons Autism Foundation, Cancer Council of Victoria, Stanley Medical Research Foundation, Medical Benefits Fund, National Health and Medical Research Council, Medical Research Futures Fund, Beyond Blue, Rotary Health, A2 milk company, Meat and Livestock Board, Woolworths, Avant and the Harry Windsor Foundation, has been a speaker for Abbot, Astra Zeneca, Janssen and Janssen, Lundbeck and Merck and served as a consultant to Allergan, Astra Zeneca, Bioadvantex, Bionomics, Collaborative Medicinal Development, Janssen and Janssen, Lundbeck Merck, Pfizer and Servier – all unrelated to this work. AT has received honoraria for lectures for Otsuka Pty, Janssen and Janssen and Servier and has served on an advisory board for Servier - all unrelated to this work. BN is supported by a NHMRC Senior Research Fellowship (1137687) and a University of Melbourne Dame Kate Campbell Fellowship. CP was supported by a NHMRC Senior Principal Research Fellowship (1105825), and NHMRC L3 Investigator Grant (1196508). Over the last three years, CP has received funding from NHMRC, Lundbeck Foundation, MRFF, and has received honoraria for talks at educational meetings and as a member of an advisory board for Lundbeck, Australia Pty Ltd.

---

**Supplementary Table 1.** Summary statistics of the cognitive measures at baseline

|  | Group | Mean | SD | n | P-values for the comparison of the 3 groups* |  |  |  |  | Effect sizes (Cohen's *d*) |  |  |
| --- | --- | --- | --- | --- | --- | --- | --- | --- | --- | --- | --- | --- |
|  |  |  |  |  | P | F | HC vs<br>PLACEBO | HC vs<br>MEDICATION | PLACEBO vs<br>MEDICATION | HC vs<br>PLACEBO | HC vs<br>MEDICATION | PLACEBO vs<br>MEDICATION |
| Digit Span:<br>Forward | HC | 10.1 | 1.9 | 42 | 0.182 | 1.73 | 0.187 | 0.080 | 0.661 | 0.30 | 0.40 | 0.10 |
|  | PLACEBO | 9.5 | 2.1 | 37 |  |  |  |  |  |  |  |  |
|  | MEDICATION | 9.3 | 2.0 | 36 |  |  |  |  |  |  |  |  |
| Digit Span:<br>Backward | HC | 7.8 | 1.9 | 42 | 0.001 | 7.98 | 0.001 | 0.001 | 0.986 | 0.78 | 0.77 | <0.01 |
|  | PLACEBO | 6.3 | 2.4 | 37 |  |  |  |  |  |  |  |  |
|  | MEDICATION | 6.3 | 1.6 | 36 |  |  |  |  |  |  |  |  |
| Digit Symbol-Coding | HC | 80.4 | 16.4 | 42 | <0.001 | 8.45 | 0.001 | 0.001 | 0.964 | 0.81 | 0.83 | 0.01 |
|  | PLACEBO | 67.1 | 16.5 | 33 |  |  |  |  |  |  |  |  |
|  | MEDICATION | 66.9 | 16.1 | 30 |  |  |  |  |  |  |  |  |
| Stroop:<br>words | HC | 99.1 | 13.8 | 42 | 0.080 | 2.59 | 0.085 | 0.038 | 0.719 | 0.40 | 0.48 | 0.09 |
|  | PLACEBO | 91.7 | 20.9 | 36 |  |  |  |  |  |  |  |  |
|  | MEDICATION | 90.1 | 21.5 | 35 |  |  |  |  |  |  |  |  |
| Stroop:<br>colours | HC | 75.9 | 13.6 | 42 | 0.007 | 5.21 | 0.006 | 0.008 | 0.952 | 0.64 | 0.62 | 0.01 |
|  | PLACEBO | 67.1 | 13.0 | 36 |  |  |  |  |  |  |  |  |
|  | MEDICATION | 67.3 | 15.4 | 35 |  |  |  |  |  |  |  |  |
| Stroop:<br>colour-word | HC | 50.1 | 11.0 | 42 | 0.002 | 6.38 | 0.001 | 0.047 | 0.147 | 0.81 | 0.46 | 0.35 |
|  | PLACEBO | 40.8 | 10.8 | 36 |  |  |  |  |  |  |  |  |
|  | MEDICATION | 44.8 | 13.0 | 35 |  |  |  |  |  |  |  |  |
| Letter<br>fluency | HC | 41.4 | 11.5 | 42 | 0.003 | 6.02 | 0.001 | 0.022 | 0.323 | 0.76 | 0.53 | 0.23 |
|  | PLACEBO | 32.8 | 10.7 | 36 |  |  |  |  |  |  |  |  |
|  | MEDICATION | 35.4 | 11.7 | 36 |  |  |  |  |  |  |  |  |
| Animal<br>fluency | HC | 23.5 | 4.1 | 42 | 0.027 | 3.73 | 0.016 | 0.030 | 0.822 | 0.56 | 0.50 | 0.05 |
|  | PLACEBO | 20.4 | 6.9 | 36 |  |  |  |  |  |  |  |  |
|  | MEDICATION | 20.7 | 5.6 | 35 |  |  |  |  |  |  |  |  |
| Immediate paired-associate recall (Trial 1) | HC | 4.2 | 2.2 | 42 | <0.001 | 9.36 | <0.001 | 0.004 | 0.252 | 0.94 | 0.67 | 0.27 |
|  | PLACEBO | 2.2 | 2.0 | 37 |  |  |  |  |  |  |  |  |
|  | MEDICATION | 2.8 | 2.3 | 36 |  |  |  |  |  |  |  |  |
| Total paired-associate learning (Trial 1-3) | HC | 17.5 | 5.1 | 42 | <0.001 | 11.72 | <0.001 | 0.001 | 0.277 | 1.04 | 0.78 | 0.26 |
|  | PLACEBO | 11.6 | 5.9 | 37 |  |  |  |  |  |  |  |  |
|  | MEDICATION | 13.1 | 5.8 | 36 |  |  |  |  |  |  |  |  |
| Delayed cued recall | HC | 6.9 | 1.6 | 42 | <0.001 | 12.80 | <0.001 | 0.001 | 0.215 | 1.10 | 0.81 | 0.29 |
|  | PLACEBO | 4.8 | 2.2 | 36 |  |  |  |  |  |  |  |  |
|  | MEDICATION | 5.3 | 1.9 | 36 |  |  |  |  |  |  |  |  |

\*Overall comparison: ANOVA; pairwise comparison: Fisher's LSD test; grey shade indicates significant result; Cohen's $d = 0.2$ is considered a 'small' effect size, 0.5 is considered a 'medium' effect size and 0.8 a 'large' effect size.

**Supplementary Figure 1.**
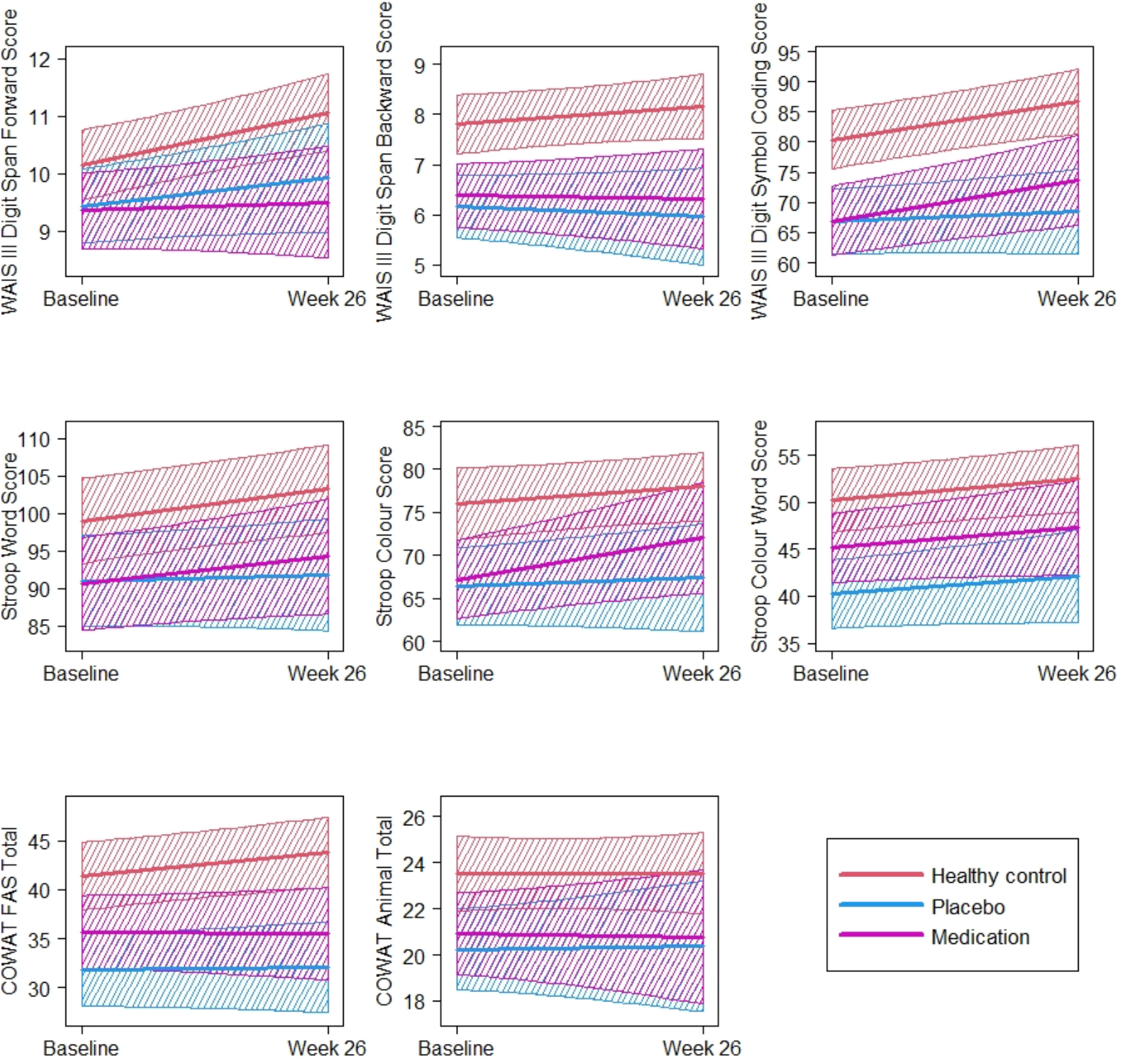
Plots of estimated trends from baseline to 6-months and associated 95% confidence bands for remaining cognitive measures.

**Supplementary Table 2.** Results of LME analysis comparing the three groups in terms of the rate of change from baseline to 6-months with patients restricted to trial completers only.

|  | P-values |  |  | Estimated rate of change |  | n |
| --- | --- | --- | --- | --- | --- | --- |
|  | Group x time interaction | Group | Time | Coefficient | Standard Error |  |
| Digit Span: forward | 0.825 | 0.212 | <0.001 | 0.033 | 0.008 | 69 |
| Digit Span: backward | 0.672 | 0.003 | 0.037 | 0.014 | 0.007 | 69 |
| Digit Symbol-Coding | 0.016 | 0.034 | <0.001 | 0.148 | 0.040 | 65 |
| Stroop: words | 0.273 | 0.200 | 0.080 | 0.085 | 0.048 | 69 |
| Stroop: colours | 0.714 | 0.199 | 0.009 | 0.097 | 0.036 | 69 |
| Stroop: colour-word | 0.254 | 0.049 | 0.017 | 0.069 | 0.029 | 69 |
| Letter fluency | 0.350 | 0.076 | 0.007 | 0.081 | 0.030 | 69 |
| Animal fluency | 0.547 | 0.275 | 0.591 | 0.012 | 0.022 | 69 |
| Immediate paired-associate recall (Trial 1) | 0.059 | <0.001 | 0.038 | 0.022 | 0.011 | 69 |
| Total paired-associate learning (Trials 1-3) | 0.035 | 0.003 | 0.213 | 0.026 | 0.021 | 69 |
| Delayed cued recall | <0.001 | 0.003 | 0.822 | 0.002 | 0.008 | 69 |

**Supplementary Figure 2.**
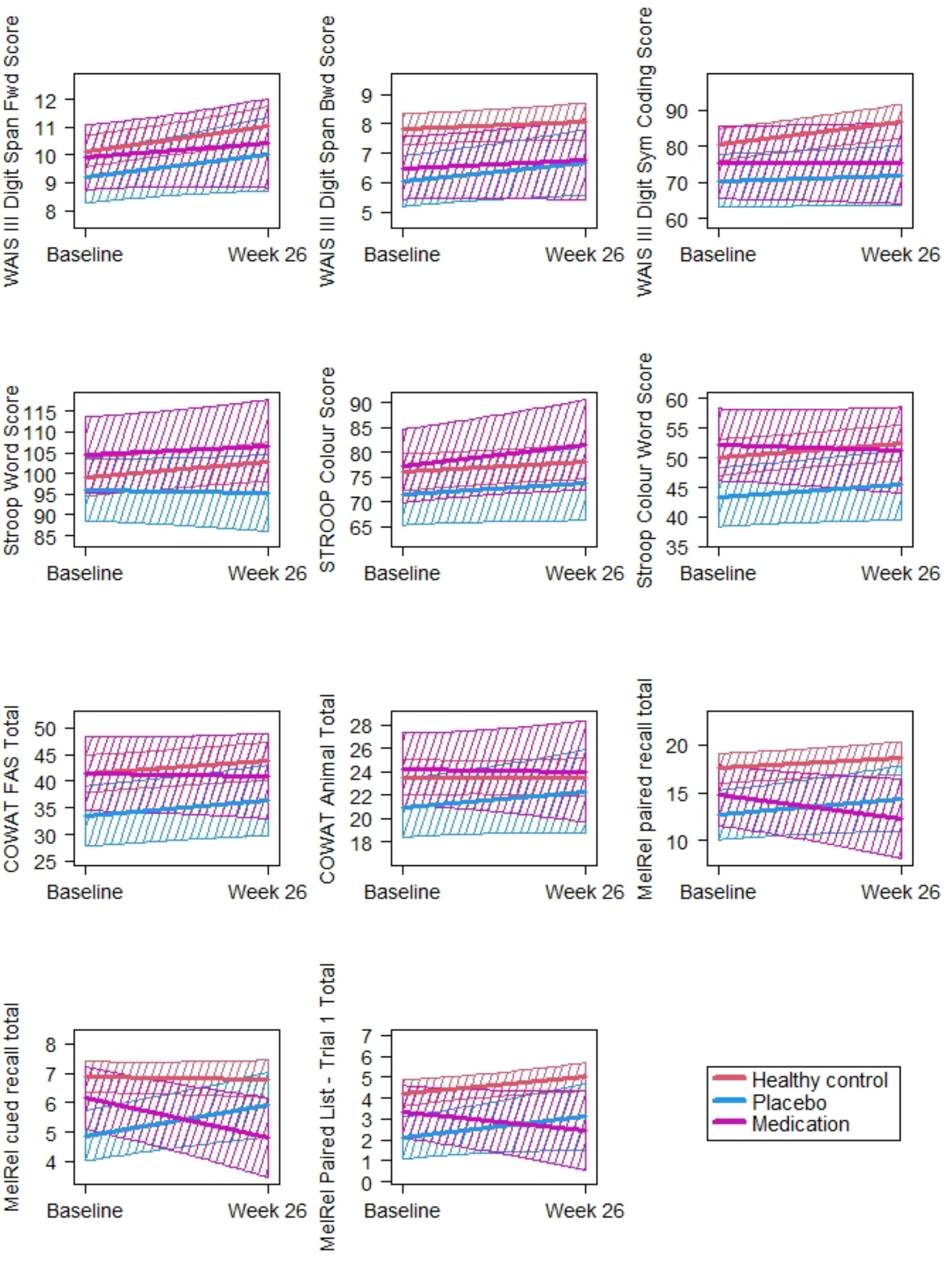
Plots of estimated trends from baseline to 6-months and associated 95% confidence bands with FEP patients restricted to trial completers only.

**Supplementary Table 3.** Results of LME analysis comparing the placebo and medication (FEP) groups in terms of rate of cognitive change from baseline to 24-month assessment.

|  | P-values |  |  | Estimated rate of change |  | n |
| --- | --- | --- | --- | --- | --- | --- |
|  | Group x time interaction | Group | Time | Coefficient | Standard Error |  |
| Digit Span: forward | 0.211 | 0.892 | 0.017 | 0.006 | 0.002 | 78 |
| Digit Span: backward | 0.554 | 0.436 | 0.021 | 0.005 | 0.002 | 78 |
| Digit Symbol-Coding | 0.252 | 0.519 | <0.001 | 0.097 | 0.017 | 69 |
| Stroop: words | 0.181 | 0.460 | 0.004 | 0.042 | 0.015 | 77 |
| Stroop: colours | 0.032 | 0.193 | <0.001 | 0.055 | 0.014 | 77 |
| Stroop: colour-word | 0.447 | 0.132 | <0.001 | 0.046 | 0.009 | 77 |
| Letter fluency | 0.670 | 0.145 | <0.001 | 0.048 | 0.011 | 78 |
| Animal fluency | 0.548 | 0.621 | 0.158 | 0.008 | 0.006 | 77 |
| Immediate paired-associate recall (Trial 1) | 0.325 | 0.964 | 0.018 | 0.006 | 0.003 | 78 |
| Total paired-associate learning (Trials 1-3) | 0.399 | 0.732 | 0.001 | 0.021 | 0.006 | 78 |
| Delayed cued recall | 0.206 | 0.902 | 0.010 | 0.006 | 0.002 | 77 |

**Supplementary Figure 3.**
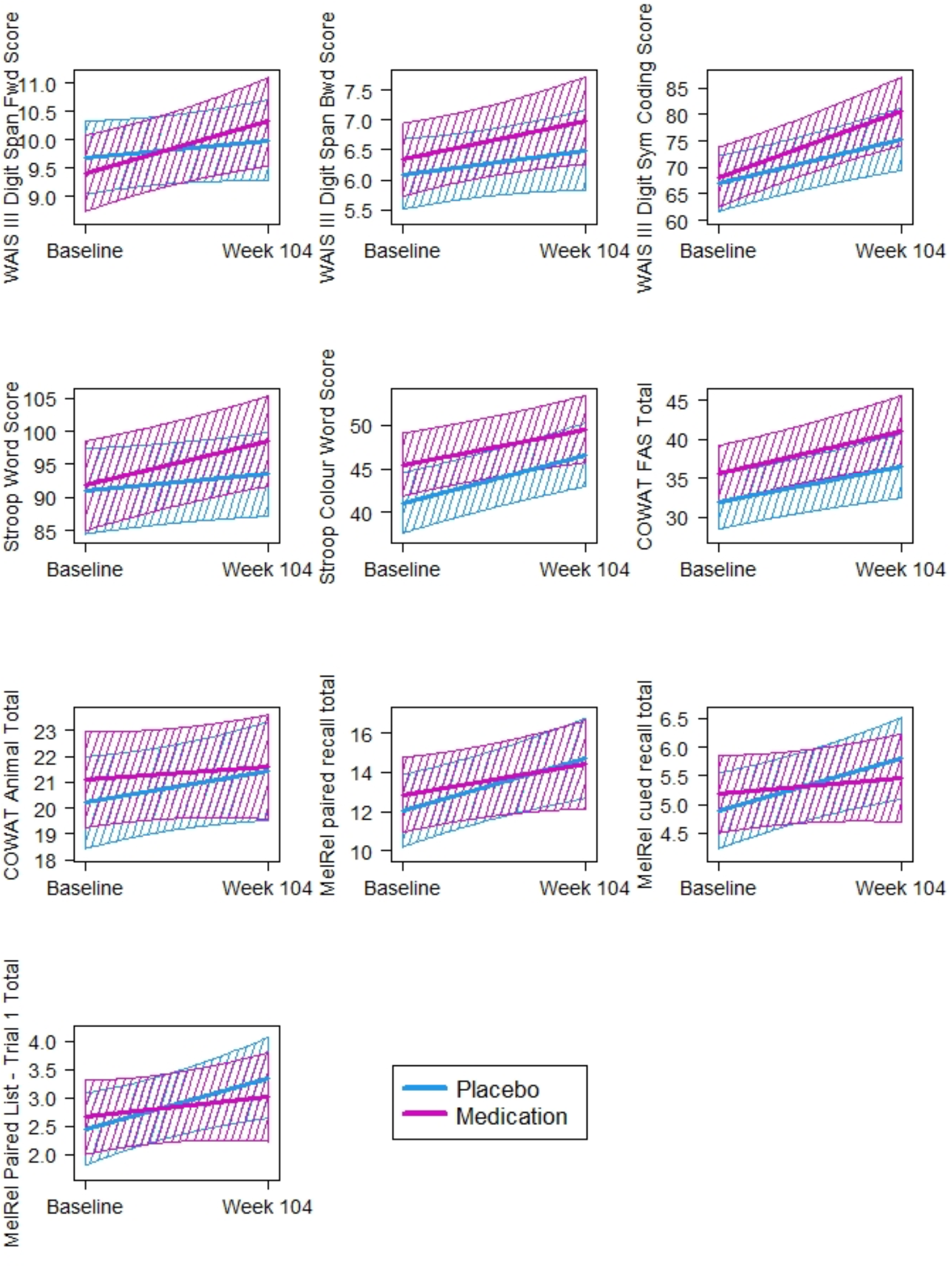
Plots of estimated trends from baseline to 24-months and associated 95% confidence bands.

**Supplementary Table 4.** Results of LME analysis comparing the placebo and medication (FEP) groups in terms of the rate of change from baseline to 24-months with patients restricted to trial completers only.

|  | P-values |  |  | Estimated rate of change |  | n |
| --- | --- | --- | --- | --- | --- | --- |
|  | Group by time interaction | Group | Time | Coefficient | S.E. |  |
| Digit Span: forward | 0.314 | 0.160 | 0.025 | 0.008 | 0.003 | 27 |
| Digit Span: backward | 0.997 | 0.720 | 0.013 | 0.010 | 0.004 | 27 |
| Digit Symbol-Coding | 0.957 | 0.352 | <0.001 | 0.096 | 0.022 | 24 |
| Stroop: words | 0.314 | 0.044 | 0.879 | 0.004 | 0.023 | 27 |
| Stroop: colours | 0.125 | 0.046 | 0.052 | 0.037 | 0.019 | 27 |
| Stroop: colour-word | 0.677 | 0.020 | 0.024 | 0.039 | 0.016 | 27 |
| Letter fluency | 0.597 | 0.132 | <0.001 | 0.065 | 0.015 | 27 |
| Animal fluency | 0.597 | 0.128 | 0.608 | 0.004 | 0.009 | 27 |
| Immediate paired-associate recall (Trial 1) | 0.182 | 0.982 | 0.563 | 0.002 | 0.004 | 27 |
| Total paired-associate learning (Trials 1-3) | 0.301 | 0.873 | 0.161 | 0.012 | 0.008 | 27 |
| Delayed cued recall | 0.163 | 0.740 | 0.327 | 0.004 | 0.004 | 27 |

## Notes

### Competing Interest Statement

The authors have declared no competing interest.

### Clinical Trial

ACTRN12607000608460

### Author Declarations

The Melbourne Health Human Research Ethics Committee approved the study (MH-HREC: #2007:616)

